# Identification of *de novo* mutations in prenatal neurodevelopment-associated genes in schizophrenia in two Han Chinese patient-sibling family-based cohorts

**DOI:** 10.1101/19011007

**Authors:** Shan Jiang, Daizhan Zhou, Yin-Ying Wang, Peilin Jia, Chunling Wan, Xingwang Li, Guang He, Dongmei Cao, Xiaoqian Jiang, Kenneth S. Kendler, Ming Tsuang, Travis Mize, Jain-Shing Wu, Yimei Lu, Lin He, Jingchun Chen, Zhongming Zhao, Xiangning Chen

**Author notes:** These authors contributed equally (Shan Jiang, Daizhan Zhou). Correspondence: Lin He or Jingchun Chen or Zhongming Zhao or Xiangning Chen.

## Abstract

Schizophrenia (SCZ) is a severe psychiatric disorder with a strong genetic component. High heritability of SCZ suggests a major role for transmitted genetic variants. Furthermore, SCZ is also associated with a marked reduction in fecundity, leading to the hypothesis that alleles with large effects on risk might often occur de novo. In this study, we conducted whole-genome sequencing for 23 families from two cohorts with matched unaffected siblings and parents. Two nonsense *de novo* mutations (DNMs) in *GJC1* and *HIST1H2AD* were identified in SCZ patients. Ten genes (*DPYSL2, NBPF1, SDK1, ZNF595, ZNF718, GCNT2, SNX9, AACS, KCNQ1* and *MSI2*) were found to carry more DNMs in SCZ patients than their unaffected siblings by burden test. Expression analyses indicated that these DNM implicated genes showed significantly higher expression in prefrontal cortex in prenatal stage. The DNM in the *GJC1* gene is highly likely a loss function mutation (pLI = 0.94), leading to the dysregulation of ion channel in the glutamatergic excitatory neurons. Analysis of rare variants in independent exome sequencing dataset indicates that *GJC1* has significantly more rare variants in SCZ patients than in unaffected controls. Data from genome-wide association studies suggested that common variants in the *GJC1* gene may be associated with SCZ and SCZ-related traits. Genes co-expressed with *GJC1* are involved in SCZ, SCZ-associated pathways and drug targets. These evidence suggest that *GJC1* may be a risk gene for SCZ and its function may be involved in prenatal and early neurodevelopment, a vulnerable period for developmental disorders such as SCZ.

## Introduction

Schizophrenia (SCZ) is a severe psychiatric disorder that profoundly affects cognitive, behavior and emotional processes, yet its etiology and pathophysiology are still largely unknown. The high heritability of SCZ suggests that genetic risk factors contribute to a significant proportion of the etiology ^1, 2^. However, the marked reduction in fecundity in SCZ patients suggests the removal of risk variants with the largest effects from the population by natural selection. Thus these variants often occur *de novo*. Indeed, the strongest genetic risk factors for SCZ identified so far are *de novo* large copy number variants (CNV) ^3^.

The availability of next-generation sequencing permits the detection of *de novo* mutation (DNM) events at the genome level. Through large-scale sequencing in parent-offspring trios, DNM have been increasingly discovered from an array of severe neurodevelopmental disorders, including Autism Spectrum Disorders (ASD) ^4^, Attention-Deficit Hyperactivity Disorder (ADHD) ^5^ and epileptic encephalopathy ^6^. For SCZ, Xu *et al*. showed a large excess of *de novo* nonsynonymous changes and DNMs presented with greater potential to affect protein structures and functions in SCZ patients ^7^; Girard *et al*. reported increased exonic DNM rate in SCZ patients ^8^; and Fromer *et al*. found that DNMs in SCZ implicated synaptic networks and DNM-affected genes in SCZ overlapped with those mutated in other neurodevelopmental disorders ^9^. Whole exome sequencing (WES) was applied predominantly in DNM identification thus far, however, few DNM studies have been based on whole genome sequencing (WGS) as WES remains a cost-effective strategy.

However, only 1.22% of DNMs are within exonic regions ^10^ and meaningful mutations may occur outside of exons, such as in regulatory elements (i.e. transcriptional promoters, enhancers and suppressors) thereby altering expression level of governed genes. Similarly, mutations within exon-intron junction regions may influence splice sites and thus lead to inappropriate expression of particular isoforms ^11^. Takata *et al*. reported that cis-acting splicing quantitative trait loci from prefrontal cortices of human were in linkage disequilibrium with SCZ genome-wide association study (GWAS) loci and DNMs near splicing sites were enriched in SCZ patients as compared to controls ^12, 13^. Emerging roles of non-coding RNAs such as microRNA, circRNA and lncRNA further call for attention to explore DNMs in non-coding regions.

We therefore conducted WGS on two cohorts of Chinese families with matched patient-sibling to capture all classes of DNMs and to more fully describe the genetic architecture of SCZ. This study was conducted to identify potential DNMs in SCZ in the Asian population, adding to the growing body of information regarding ethnicity-specific DNMs.

## Methods

### Subjects

Subjects were drawn from two distinct cohorts, Taiwan and Shanghai, of Han Chinese origin. Recruitment of subjects from the Taiwan cohort was described in previous publications ^14, 15^. The rationale for combining the two cohorts was to increase the power to detect DNMs because each of the two cohorts has small sample size but they are ethnically homogeneous. Briefly, families with at least three siblings, two of whom were diagnosed with SCZ, were recruited in the Taiwan Schizophrenia Linkage Study (TSLS) from 1998 to 2002. All recruited subjects were interviewed using the Diagnostic Interview for Genetic Studies (DIGS) ^16^, accompanied with the Family Diagnostic Interview for Genetic Studies (FIGS) (https://www.nimhgenetics.org/resources/clinical-instruments/figs/list-of-figs). Final diagnostic assessment was based on the criteria of the fourth edition of the Diagnostic and Statistical Manual (DSM-IV), joined with the record of DIGS, FIGS, interviewer notes, and hospital anamnesis. For the Shanghai cohort, families from the Bio-x SCZ Biobank with at least three siblings, two of whom were diagnosed with SCZ, were selected. All families from the Bio-x SCZ Biobank were recruited from the city of Shanghai and the provinces of Hebei, Liaoning and Guangxi from 2001 to 2003. All individuals with SCZ were interviewed by two independent psychiatrists and diagnosed according to DSM-IV criteria. A total of 23 families (10 from Taiwan and 13 from Shanghai), with SCZ patients, matched unaffected siblings and parents, were used in this study (Figure 1). For detailed demographic characteristics of all individuals in the 23 families, please refer to Supplementary Table S1. All subjects gave written informed consent with the approval of the local research ethics committees.

**Figure 1.**
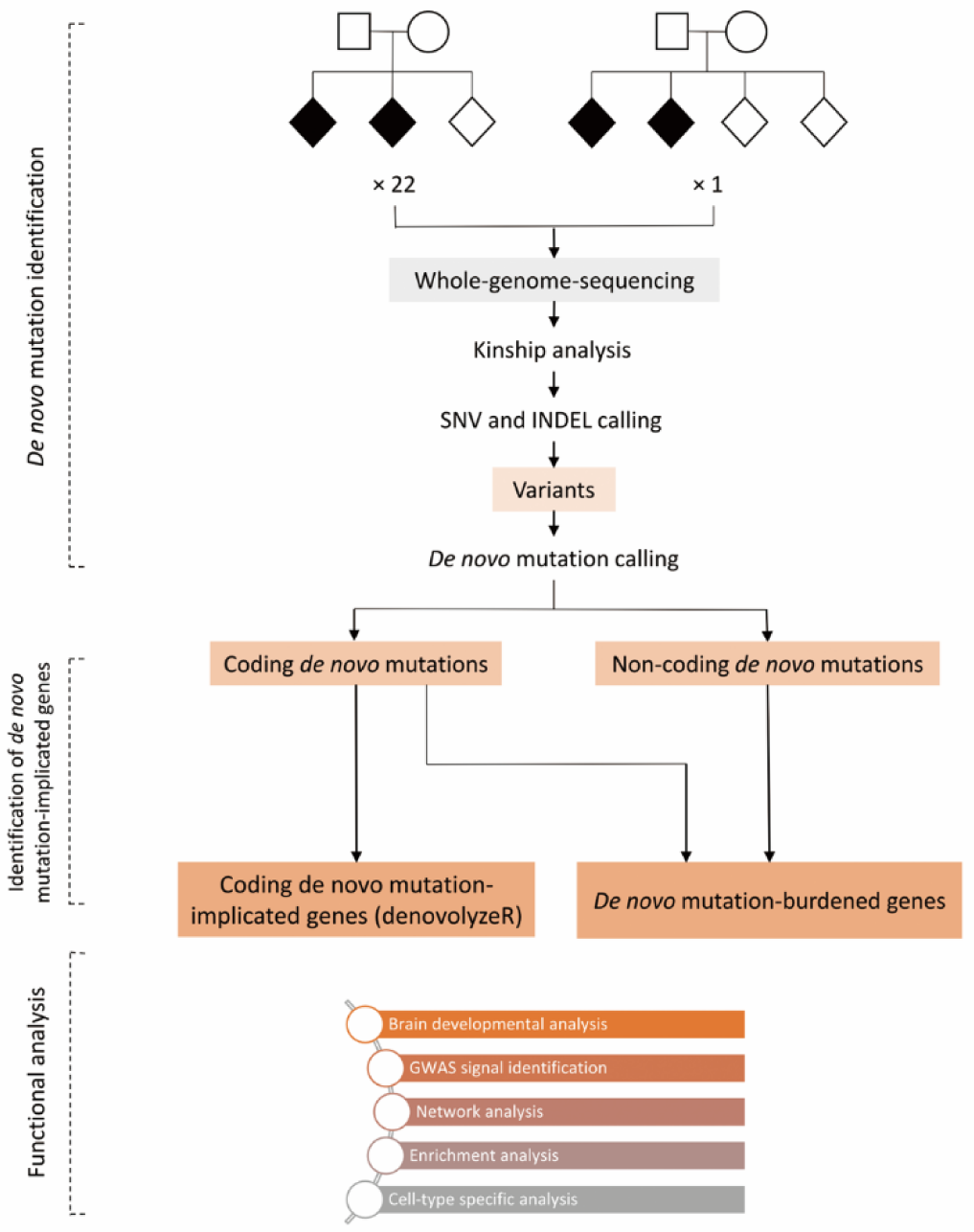
Schematic of genetic data processing, DNM identification and functional analysis in 23 families with schizophrenia patients and matched unaffected siblings. SNV, single nucleotide variant; INDEL, insertion and deletion.

### Whole genome sequencing

For the subjects of Taiwan cohort, whole blood samples were collected with anticoagulant (EDTA) tubes and sent to the National Institute of Mental Health (NIMH) Repository and Genomics Resource (RGR). Lymphocytes from the whole blood samples were transformed into immortalized lymphoblastoid cell lines and stored. The DNA samples extracted from the cell lines were used for WGS. WGS was carried out on the Illumina HiSeq 2000 platform using paired-end chemistry with 75 base-pair read length through NovoGene, Inc. (Beijing, China). For detailed description please refer to previous publication ^17^. WGS data for the subjects of Taiwan cohort can be accessed in BioProject of NCBI: https://www.ncbi.nlm.nih.gov/bioproject/PRJNA551447. For the subjects from Shanghai cohort, whole blood samples were also collected with anticoagulant tubes. DNA was extracted from blood lymphocytes by standard procedures using FlexiGene DNA kits (Fuji, Tokyo, Japan). DNA libraries were prepared using protocols recommended by Illumina (Illumina, San Diego, CA). WGS was performed on Illumina HiSeq-X Ten platform with 150 base-pair read length through Cloud Health Genomics Ltd. (Shanghai, China).

### Quality control and variant calling

FastQC (v0.11.8) was used to perform quality checks on all samples across Taiwan and Shanghai cohorts (Supplementary Figure S1).

The GATK best practices of variant calling were applied to process all raw reads from both Taiwan and Shanghai cohorts ^18^. Raw sequencing reads in FASTQ format were aligned to the GRCh37 build of the human reference genome with BWA-mem ^19^. Then the aligned reads in BAM format were sorted, indexed and marked with duplicate reads with Picard Tools. Reads containing indels were realigned with GATK’s IndelRealigner tool. Next, GATK was used to perform Base Quality Score Recalibration (BQSR). Quality control after alignment was performed using Picard (v2.20.4) CollectAlignmentSummaryMetrics (Supplementary Table S2). Variant calling was performed across all samples with GATK (v4.1.1.0) HaplotypeCaller. For detailed variant calling pipeline, please refer to Supplementary Figure S2. To further systematically remove potentially false positively called variants, we applied the following quality filters for each variant: (1) the quality score normalized by allele depth is less than 2; (2) the root mean square of the mapping quality is less than 40; (3) the strand bias is more than 60; (4) three consecutive variants were clustered within 10 bases; and (5) the FILTER tag is not PASS.

### Kinship analysis, DNM calling and annotation

We used PLINK to perform kinship analysis ^20^. Briefly, for a given family, if the father was shown to be within the third degree relative to the mother, the family would be excluded; if the child was not shown to be the first degree relative to the parents, the child would be excluded.

To ensure the DNM calls with high confidence, three tools, GATK PhyseByTransmission (PBT), TrioDeNovo (v0.0.6) and DeNovoGear (v.develop), were used to evaluate the calls ^21-23^. Only DNMs called by all three tools consistently were considered as candidate DNMs.

PBT was run with the following command:

~~~
java –jar GenomeAnalysisTK.jar -T PhaseByTransmission -R human_g1k_v37_decoy.fasta –V parent_child_trio.vcf -prior 1.0e-8 -mvf PBT_result.vcf
~~~

TrioDeNovo was run with the following command:

~~~
triodenovo --ped parent_child_pedigree.ped --in_vcf parent_child_trio.vcf --mu 1.0e-8 --out_vcf TrioDeNovo_result.vcf
~~~

DeNovoGear was run with the following command:

~~~
dng dnm --ped parent_child_pedigree.ped –vcf parent_child_trio.vcf -s 1.0e-8 –write DeNovoGear_result.vcf
~~~

After obtaining consistently called DNMs, the following five criteria were applied to retain high quality calls tagged with PASS in the FILTER annotation: (1) quality score is greater than or equal to 30; (2) genotypes of the parents are homozygotes; (3) genotype of the child is heterozygote; (4) phred-scaled maximum likelihood of heterozygote for the parents is less than 50; and (5) phred-scaled maximum likelihood of heterozygote of the child is 0. DNMs with MAF greater than 0.01 in East Asian populations were further excluded by querying gnomAD (https://gnomad.broadinstitute.org/) and EXac (http://exac.broadinstitute.org/). Please refer to Supplementary Figure S2 for detailed pipeline of DNM calling and Supplementary Figure S3 for the distributions of DNM quality scores. Remaining DNMs were annotated using ANNOVAR ^24^.

### Polymerase chain reaction (PCR)-based Sanger sequencing validation

For the families where candidate DNMs were found, DNA from all members of the family was subjected to PCR-based Sanger sequencing by capillary electrophoresis according to standard molecular biology practices (ABI 3130 genetic analyzer, ThermoFisher Scientific). Primer3Plus was used to design the PCR primers ^25^. For the *GJC1* mutation, the forward primer sequence was 5’-TTAGGTTTGGGTTGGCTCTG-3’ and the reverse primer sequence was 5’-CACGGTGAAGCAGACAAGAA-3’. For the *HIST1H2AD* insertion, the forward primer sequence was 5’-CTCGTTTTACTTGCCCTTGG-3’ and the reverse primer sequence was 5’-ACAACAAGAAGACCCGCATC-3’.

Reactions were performed on an Eppendorf MasterCycler (Eppendorf North America, New York, USA) under the following cycling conditions: denaturation at 95 °C for 3 min, 35 cycles of 95LJ°C for 15LJsec, 55 °C for 20 sec, 72 °C for 30 sec, and a final extension at 72 °C for 5 min. Sanger sequencing data was then analyzed using Chromas software (https://technelysium.com.au/wp/).

### Identification of genes implicated by coding DNMs in SCZ

As the spontaneous background mutation rates vary greatly between genes, those carrying relatively more protein-coding DNMs in cases are not necessarily implicated by DNMs ^26^. To identify those genes implicated by protein-coding DNMs or of which the DNMs occurred higher than the background mutation rates, the R package denovolyzeR was used to analyze protein-coding DNMs based on a mutation model developed previously ^26^. Briefly, denovolyzeR estimates underlying mutation rate based on trinucleotide context and incorporates exome depth and divergence adjustments based on macaque-human comparisons over a ± 1-Mb window and accommodates known mutational biases, such as CpG hotspots. By applying the underlying mutation rate estimates, denovolyzeR generates prior probabilities for observing a specific number and class of mutations (synonymous, missense, nonsense, splice-site and frameshift) for a given gene.

To validate whether the genes implicated by coding DNMs were also implicated by rare variants, the population-based SCZ Swedish case-control cohort with whole exome sequences and variants called were used to perform the analysis ^27^. Variants with MAF greater than 0.01 were excluded and only rare variants were retained subsequently. The association between the set of rare variants from Swedish case-control cohort located in the exons of a given gene implicated by coding DNM and phenotype was tested using SKAT ^28^.

### DNM burden test

To determine whether some genes carry more DNMs in SCZ patients than expected by chance, we performed the DNM burden test for each gene potentially implicated by DNM. Human brain-specific gene enhancer information was included from PsychENCODE (http://resource.psychencode.org/) and human-specific gene promoter information from Eukaryotic Promoter Database (EPD, https://epd.epfl.ch//index.php) ^29, 30^. For a given gene, DNMs occurring in its gene body, which includes both exons and introns, brain-specific gene enhancer and gene promoter regions were considered as burdens. For a given gene, we compared the number of DNMs mapped to these regions in SCZ patients with the number of DNMs mapped to these regions in unaffected siblings, and then assessed the significance of the comparison using 10,000 within-sibship case-control label-swapping permutations. *P* value was calculated as the proportion of permutations with relative risk (RR) as or more extreme than in the observed data.

### Developmental expression of DNM-implicated genes

Multiple lines of evidence have shown that prenatal maternal infection, malnutrition and stress are risk factors for SCZ ^31-33^. The neurodevelopmental model of SCZ posits that a perturbation in early brain development leads to an altered brain developmental trajectory that is sensitive to molecular changes associated with development and environmental experience, consequently converging on the emergence of SCZ in early adulthood ^34^. It was hypothesized that DNMs drove dysfunction of genes in early brain development and that this dysfunction confers risks for subsequent SCZ. To determine whether DNM-implicated genes (loss-of-function DNM genes and DNM-burdened genes) were involved in early brain development, human brain developmental expression data from BrainCloud and BrainSpan was evaluated ^35, 36^. The expression data of BrainCloud consisted of data derived from the prefrontal cortices of 269 individuals ^35^, whereas the expression data of BrainSpan consisted of 42 brain specimens across 13 developmental states in 8-16 brain structures ^36^. BrainCloud expression data was examined by comparing the expression of a DNM gene in prenatal stage to the expression in postnatal stage and BrainSpan expression data was evaluated by leveraging the developmental effect scores curated in a previous publication ^37^. A developmental effect score measures the effect of age on expression per gene per brain structure, with a higher developmental effect score of a gene in a given brain structure indicating a stronger involvement of brain development in that structure. By 10,000 gene label-swapping permutation, *P* value of a DNM gene for a given brain structure was calculated as the proportion of permutations with developmental effect scores as or more extreme than the observed value. DNM genes were then further evaluated in brain developmental expression data from other species (macaque and mouse) ^38, 39^.

### Genetic susceptibilities of DNM-implicated genes in SCZ and SCZ-related traits

To determine whether DNM-implicated genes are among the loci found by the genome-wide association study (GWAS) of SCZ or SCZ-related traits, we used GWAS summary statistics from SCZ ^1^, ASD ^40^, ADHD ^41^, bipolar disorder (BD) ^42^, major depressive disorder (MDD) ^43^, intelligence ^44^, educational attainment (EA) ^45^, cognitive performance (CP) ^45^, and smoking and drinking ^46^. The criterion for annotating a single nucleotide polymorphism (SNP) to a given gene was that the GWAS SNP is located within 10 kb of the gene boundaries.

### Co-expression and enrichment analysis

To explore the potential functions or pathological pathways affected by DNM-implicated genes, expression data from BrainCloud was leveraged by retrieving genes highly co-expressed with the candidate DNM genes. To identify enrichments in gene ontologic features, biological pathways, diseases and drug targets, WebGestalt was used (http://www.webgestalt.org/) ^47^. In WebGestalt, genes co-expressed with the candidate DNM gene were input as the target gene set and all genes in the BrainCloud expression data were input as the reference gene set. Enrichments with B-H FDR-corrected *P* values less than 0.05 were considered significantly enriched.

### Cell-type specific expression analysis

To examine cell-type specific expression of DNM-implicated genes, brain tissue single nucleus RNA-seq (snRNA-seq) data of middle temporal gyrus (MTG), of which the cells have been sub-typed from Allen Brain Atlas (https://celltypes.brain-map.org/rnaseq) and single cell expression data from PsychENCODE were utilized. Single cell expression data from PsychENCODE were merged from multiple brain regions, including frontal cortex, visual cortex and cerebellar hemisphere ^48^. Raw read count data was normalized by log-transformation using R package Seurat ^49^. For a given DNM gene, the dominant cell type(s) with high expression were determined by pair-wise Wilcoxon test.

## Results

The goal of our study was to identify DNM disturbed genes, of which the dysfunctions can contribute to the pathogenesis of SCZ. We identified coding DNM-implicated genes and genes carrying more DNM burdens in SCZ patients than their unaffected siblings. These DNM-implicated genes were potentially detrimental and subsequent analyses were performed to explore their pathological effects (Figure 1).

### Identification of DNMs and genes implicated by DNMs

To ensure parental unrelatedness and that the children are indeed biological offspring of their parents, kinship analysis was performed for each individual family. For the Taiwan cohort, one family (Family ID: 35-04560) was excluded as the father is within the third degree relative of the mother, one family (Family ID: 35-93405) was excluded as the father was not found to be related to any of the children and one child in one family was excluded as he was found to be unrelated to his parents (Individual ID: 35-02497-01) (Supplementary Table S1). Three children from Shanghai cohort were excluded as they were unrelated to their respective parents (Individual IDs: CHG000225, CHG000236 and CHG000246) (Supplementary Table S1). A total of 21 families were retained for analyses. For detailed information of the families recruited and the individuals in each family in the Taiwan and Shanghai cohorts and the families or individuals excluded due to unrelatedness, please refer to Supplementary Table S1.

In this study, 70.71 ± 6.83 *de novo* point mutations and 6.31 ± 3.64 *de novo* indel mutations with high confidence per individual were identified. The observed *de novo* point mutation rate of 1.145 × 10^−8^ was consistent with the neutral expectation of 1.140 × 10^−8^ (*P* = 0.95, two-sided exact binomial test) ^23^. The observed *de novo* indel mutation rate of 1.022 × 10^−9^ was consistent with the neutral expectation of 1.420 × 10^−9^ (*P* = 0.500, two-sided exact binomial test) ^23^. All DNMs were checked visually by Integrative Genomics Viewer (IGV). Overall, the DNMs occurred in non-coding regions predominantly in intergenic regions (Supplementary Figure S4). No obvious difference was observed between unaffected siblings and SCZ patients with regards to the distributions of DNM locations relative to genes (Supplementary Figure S4). No obvious differences were observed between unaffected siblings and SCZ patients with regards to the percentages of DNMs occurring in exons (unaffected siblings: 29.85% versus SCZ patients: 30.29%) and introns (unaffected siblings: 1.53% versus SCZ patients: 2.03%). The nonsynonymous-to-synonymous ratio in SCZ patients was not found to be different from the ratio in unaffected siblings ^50^, which might be attributed to the limited sample size. Two nonsense loss-of-function DNMs implicating *GJC1* and *HIST1H2AD* respectively were identified in SCZ patients (Table 1 and Supplementary Table S4). The two loss-of-function DNMs were visually verified by IGV (Supplementary Figure S5 and S6) and then were confirmed by Sanger sequencing (Supplementary Figure S7 and S8). To examine whether the occurrence rates of nonsense loss-of-function DNMs in *GJC1* and *HIST1H2AD* in SCZ patients were higher than expected by chance, we compared the nonsense mutation probabilities of *GJC1* and *HIST1H2AD* to those of all genes calculated in Samocha *et al*. ^51^. Nonsense DNMs were not prone to occur in DNA regions of *GJC1* and *HIST1H2AD* (Supplementary Figure S9). No loss-of-function DNMs were identified in unaffected siblings on the whole genome scale (Supplementary Table S3).

**Table 1.**
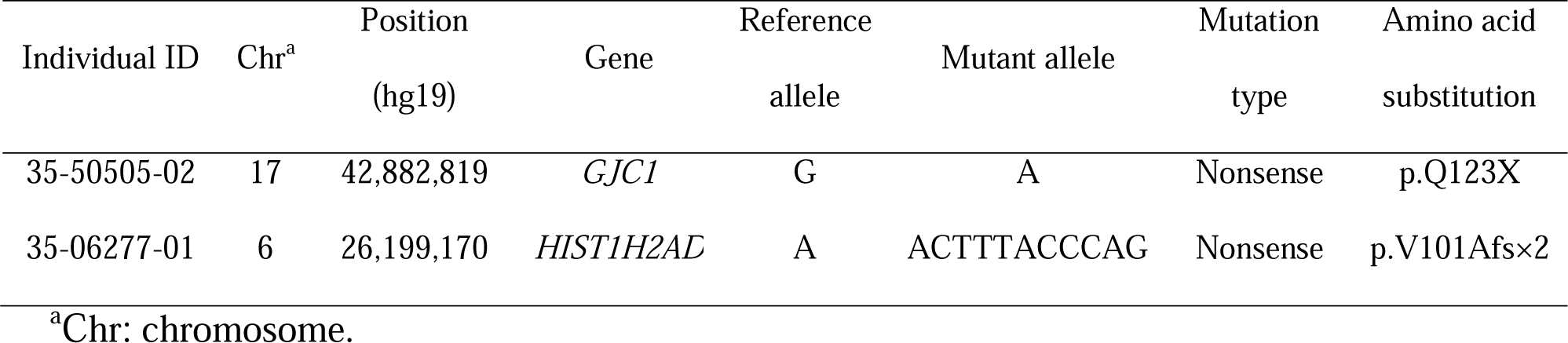
Loss-of-function DNMs identified in schizophrenia patients

Except for the two nonsense loss-of-function DNMs, we also investigated other DNMs in coding regions to find damaging DNMs because DNMs occurring in coding regions may change protein structures physically (Figure 1 and Supplementary Table S3). A detailed list of these DNMs are shown in Supplementary Table S4. To identify genes implicated by these protein-coding DNMs, denovolyzeR was used. Due to the relatively small sample size of this study, Bonferroni correction was used to reduce the false discovery rate. Only *GJC1* and *HIST1H2AD* carrying nonsense DNMs were significantly identified to be implicated by DNMs (Supplementary Table S5). We applied another background mutation rate-based model, referred to as the chimpanzee-human divergence model, which also showed *GJC1* and *HIST1H2AD* were significantly implicated by the nonsense DNMs (*P* value = 1.56 × 10^−5^ for *GJC1* and *P* value = 1.82 × 10^−5^ for *HIST1H2AD*) ^52^. Of note, the *GJC1* nonsense DNM was predicted to be extremely close to the most severe 0.1% of mutations by combined annotation dependent depletion (CADD) (Supplementary Table S4; mutations with CADD phred-like scores greater than or equal to 30 is the most severe 0.1% of mutations) ^53^. Moreover, *GJC1* falls into the haploinsufficient category with a high probability of loss-of-function intolerance (pLI) of 0.94, therefore is extremely intolerant of loss-of-function variation ^54^.

To investigate whether *GJC1* and *HIST1H2AD* were also implicated by rare variants as DNMs in SCZ, we analyzed SCZ Swedish case-control cohort comprised of 4969 SCZ patients and 6245 controls. Rare variants in the exons of *GJC1* were significantly associated to SCZ (Table 2).

**Table 2.** Rare variant association tests for the loss-of-function DNMs identified in schizophrenia patients

| Gene | Rare variant frequency in SCZ | Rare variant frequency in control | SKAT $P$ value |
| --- | --- | --- | --- |
| *GJC1* | 0.0115 | 0.0091 | $5.60 \times 10^{-3}$ |
| *HIST1H2AD* | 0.0029 | 0.0043 | $6.28 \times 10^{-1}$ |

### Identification of genes burdened with DNMs in SCZ patients

Previous studies have shown that 99% of the DNMs occurred in non-coding regions ^10^. DNMs occurring in regulatory regions can potentially disturb the bindings of transcriptional factors and thus influence gene expression. If a gene or regulatory regions of the gene carry more DNMs in SCZ patients than their unaffected siblings, this gene may have a role in disease predisposition. Based on this assumption, for a given gene, we compared the number of DNMs that occurred in brain-specific enhancer, promoter and gene body in SCZ patients to the number of DNMs in unaffected siblings by within-sibship case-control label-swapping permutation. Before calculation, to reduce the falsely identified genes by chance, we excluded genes with low total DNM counts in both SCZ patients and unaffected siblings (≤ 3). Due to the relatively small sample size, Bonferroni correction was used to reduce the false discovery rate. Ten genes, *DPYSL2, NBPF1, SDK1, ZNF595, ZNF718, GCNT2, SNX9, AACS, KCNQ1* and *MSI2*, were identified to carry more DNMs in SCZ patients than their unaffected siblings. Multiple DNMs implicated enhancers for *DPYSL2, NBPF1, SNX9* and *MSI2* (Supplementary Table S6), suggesting the dysregulation of these genes may predispose an individual to SCZ. One single enhancer can regulate the expression of multiple genes. Therefore, single DNM implicating an individual enhancer can disturb the expression of multiple genes. We leveraged transcription factor (TF)-enhancer-target gene linkage data from PsychENCODE to establish the gene regulatory network disturbed by the DNMs implicating the enhancers identified by DNM burden test (Figure 2). For detailed list of the DNMs found in the ten genes reported here, please refer to Supplementary Table S6. Interestingly, no DNM occurred in promoters of the ten genes.

**Figure 2.**
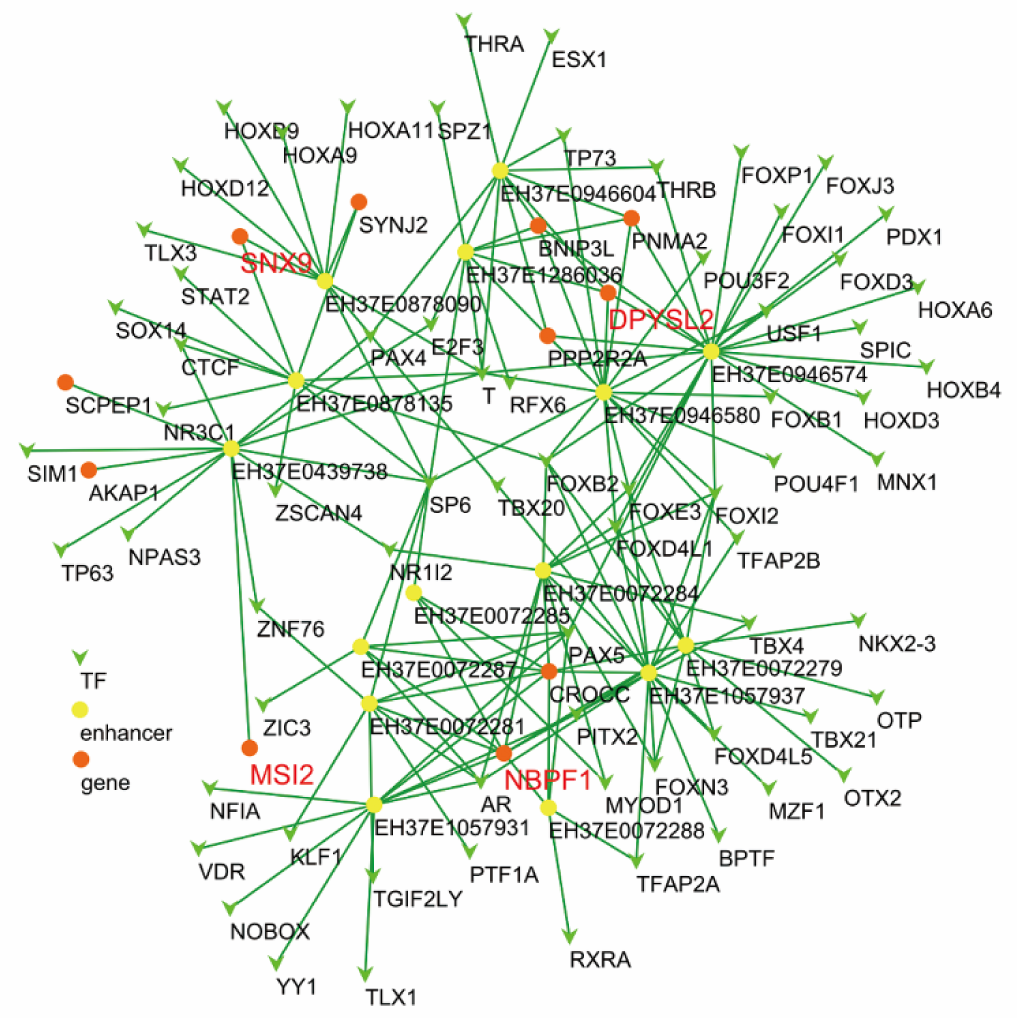
Gene regulatory network disturbed by the DNMs implicating the enhancers identified by DNM burden test. Light green arrow nodes are the transcription factors. The yellow round nodes are the enhancers perturbed by the DNMs. The Orange round nodes are the genes. The edge between transcription factor and enhancer represent the transcription factor can bind to the enhancer without the perturbation of DNM. The edge between enhancer and gene represent the enhancer can enhance the expression of the gene without the perturbation of DNM. Red labeled genes were those identified by DNM burden test.

### DNMs occurred in genes involving in early brain development

In the following analyses, coding DNM-implicated genes (*GJC1* and *HIST1H2AD*, Supplementary Table S5) and DNM-burdened genes (Table 3) were as assumed to be detrimental DNM genes to explore their potential functions that might be related to SCZ.

**Table 3.** Genes significantly enriched with higher DNM burden in schizophrenia patients than unaffected siblings

| Gene symbol | DNM count in unaffected siblings | DNM count in schizophrenia patients | *P*-value | Bonferroni<br>*P*-value |
| --- | --- | --- | --- | --- |
| *DPYSL2* | 0 | 6 | $1 \times 10^{-4}$ | $3.4 \times 10^{-3}$ |
| *NBPF1* | 0 | 6 | $1 \times 10^{-4}$ | $3.4 \times 10^{-3}$ |
| *SDK1* | 1 | 5 | $1 \times 10^{-4}$ | $3.4 \times 10^{-3}$ |
| *ZNF595* | 1 | 5 | $1 \times 10^{-4}$ | $3.4 \times 10^{-3}$ |
| *ZNF718* | 1 | 5 | $1 \times 10^{-4}$ | $3.4 \times 10^{-3}$ |
| *GCNT2* | 0 | 5 | $1 \times 10^{-4}$ | $3.4 \times 10^{-3}$ |
| *SNX9* | 0 | 5 | $1 \times 10^{-4}$ | $3.4 \times 10^{-3}$ |
| *KCNQ1* | 1 | 4 | $1 \times 10^{-4}$ | $3.4 \times 10^{-3}$ |
| *AACS* | 1 | 3 | $1 \times 10^{-4}$ | $3.4 \times 10^{-3}$ |
| *MSI2* | 1 | 3 | $1 \times 10^{-4}$ | $3.4 \times 10^{-3}$ |

Early neurodevelopmental events have been implicated in the pathogenesis of SCZ ^34^. Genes implicated in SCZ function in processes important to fetal brain development ^50, 55^. To determine whether the detrimental DNM genes, including *GJC1* and *HIST1H2AD* implicated by nonsense mutations (Supplementary Table S5) and genes carrying more DNMs in SCZ patients (Table 3), were involved in brain development, we leveraged the developmental expression data of prefrontal cortices (PFC), one of the most highly implicated brain regions in SCZ, from BrainCloud. Nine of the 13 detrimental DNM genes, including the two loss-of-function DNM-implicated genes *GJC1* and *HIST1H2AD*, and seven DNM-burdened genes *DPYSL2, NBPF1, SDK1, ZNF595, ZNF718, KCNQ1* and *SNX9*, showed biased higher expression in prenatal stage compared to that of the expression in postnatal stage (Figure 3). The large proportion of 9/12 potentially detrimental DNM genes with higher expression in prefrontal cortices was more than expected by chance (Supplementary Figure S10, *P* = 0.05, hypergeometric test). We next sought to examine whether the involvements of detrimental DNM genes in PFC development were preserved in other brain regions. The genes with available developmental data in BrainSpan were analyzed. It was found that *GJC1, SDK1* and *GCNT2* were still involved in the developments of other brain regions (Supplementary Figure S11). To examine whether the involvements of detrimental DNM genes in human brain development were conserved across species, we first queried the brain developmental expression data of macaque, a primate species evolutionarily close to *Homo sapiens*. Only DNM genes with expression data of macaque were analyzed. *GJC1, HIST1H2AD, DPYSL2, SDK1* and *MSI2* showed higher expression in prenatal stage than the expression in postnatal stage (Supplementary Figure S12). We then queried the brain developmental expression data of mouse. Since the developmental data of mouse were limited in postnatal stage, we applied linear regression to examine the change of DNM gene expression level with time. The expression levels of *GJC1, DPYSL2, SDK1* and *AACS* significantly decreased with time (Supplementary Figure 13), which implied their involvements in prenatal neurodevelopment of mouse. Specifically for *GJC1*, Leung et al. showed that *GJC1* displayed high expression in embryonic stage followed by a massive postnatal decrease in the rat midbrain-floor, where dopaminergic neurons were mostly populated ^56^.

**Figure 3.**
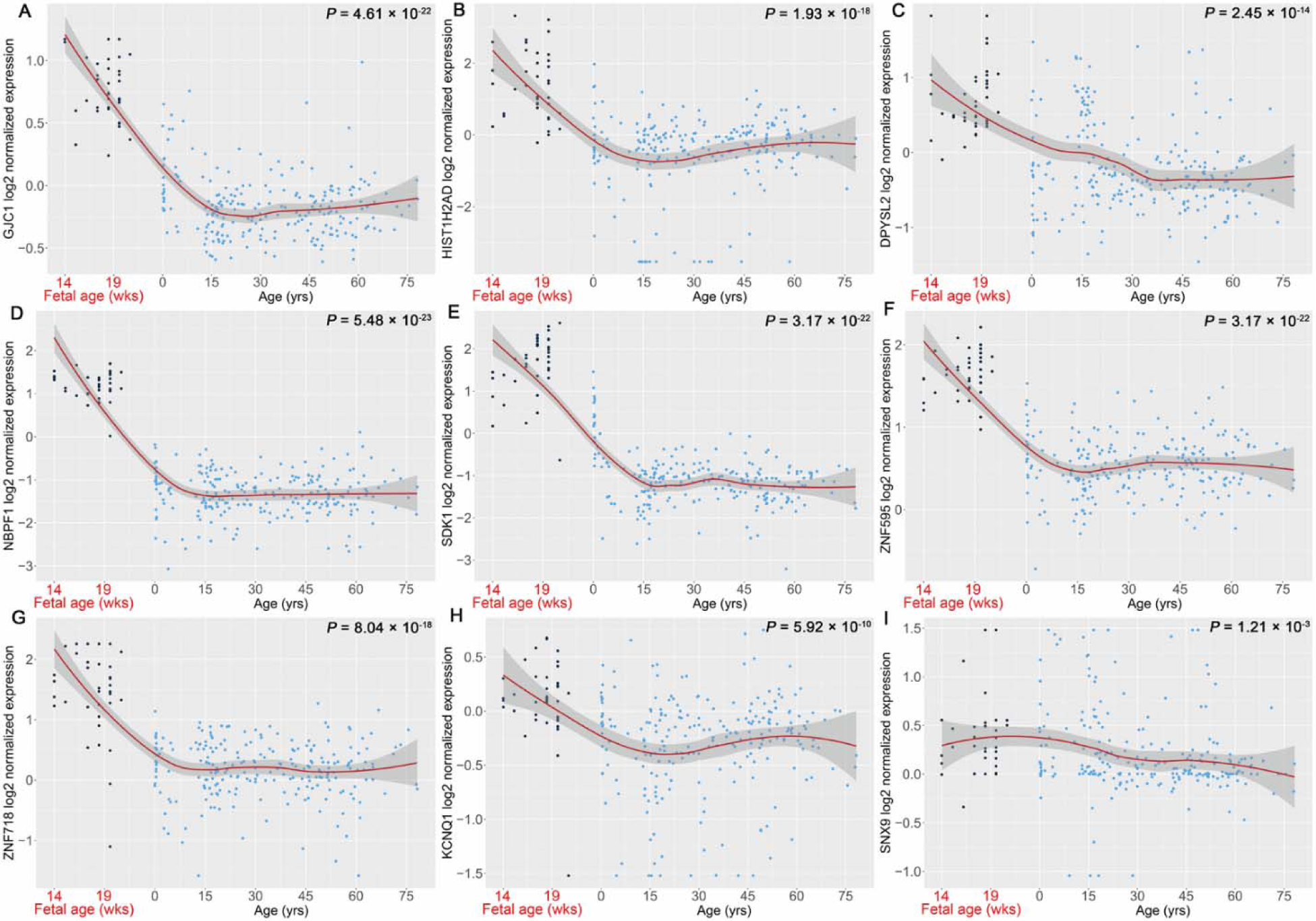
Detrimental DNM genes implicated in early brain development in PFC. A, *GJC1*; B, *HIST1H2AD*; C, *DPYSL2*; D, *NBPF1*; E, *SDK1*; F, *ZNF595*; G, *ZNF718*; H, *KCNQ1*; I, *SNX9*. Black dots represent samples from prenatal stage. Blue dots represent samples from postnatal stage. *P* values were derived from Wilcoxon rank sum test by comparing gene expression from prenatal stage to postnatal stage.

### Common variants in detrimental DNM genes may influence risks of SCZ-associated traits

To determine whether the detrimental DNM genes are involved more broadly in SCZ, we asked whether common variants present in the DNM genes confer risk for SCZ and SCZ-associated traits. We extracted all SNPs that occur within 10 kb of the DNM genes. For a given trait, the SNP with the minimum *P* value mapped to the gene was used to represent the gene risk on the trait. Genes with minimum SNP *P* values less than the suggestive threshold of 1 × 10^−5^ were considered as risk genes for the trait. We found that the DNM genes influence risk for SCZ-associated traits, including intelligence, educational attainment, cognitive performance, smoking and drinking primarily (Supplementary Table 7). Of note, *GJC1, HIST1H2AD* and *SDK1* confer risks for multiple SCZ-associated traits with SNPs passed the genome-wide significance threshold of 5 × 10^−8^.

### GJC1 co-expressed with multiple potassium channel genes and is a potentially target for SCZ

Next, we focused on the loss-of-function DNM gene *GJC1*, a member of the connexin gene family, which showed strong evidence of SCZ susceptibility (Supplementary Table S8). DNMs in other brain-associated connexin genes were also found in psychiatric patients (Supplementary Table S9). *GJC1* showed higher expression in prenatal stage than the expression in postnatal stage in PFC and multiple other brain regions, and its involvement in early brain development was conserved across species (Figures 3; Supplementary Figures S11, S12 and S13). Moreover, common variants present in *GJC1* confer risks for intelligence, educational attainment, cognitive performance and alcohol abuse, which were associated with SCZ (Supplementary Table S7) ^57-60^.

To explore the pathogenic effects of *GJC1*, we first sought to identify the genes that might be impacted by the dysfunction of *GJC1*. We performed co-expression analysis with Pearson correlation coefficient greater than 0.8 or less than −0.8 in PFC to identify these genes (Figure 4A and supplementary Table S10). Then we leveraged the genes co-expressed with *GJC1* to perform enrichment analysis to identify functions or pathways involved by *GJC1*. Interestingly, the genes co-expressed with GJC1 were enriched in SCZ, SCZ drug zuclopenthixol and SCZ-associated functions or pathways, including potassium ion transport (Figure 4B and Supplementary Table S11). Of note, multiple potassium ion channel genes were negatively co-expressed with *GJC1* (Figure 4A), suggesting their dysfunctions may be subsequent to the dysfunction of *GJC1*.

**Figure 4.**
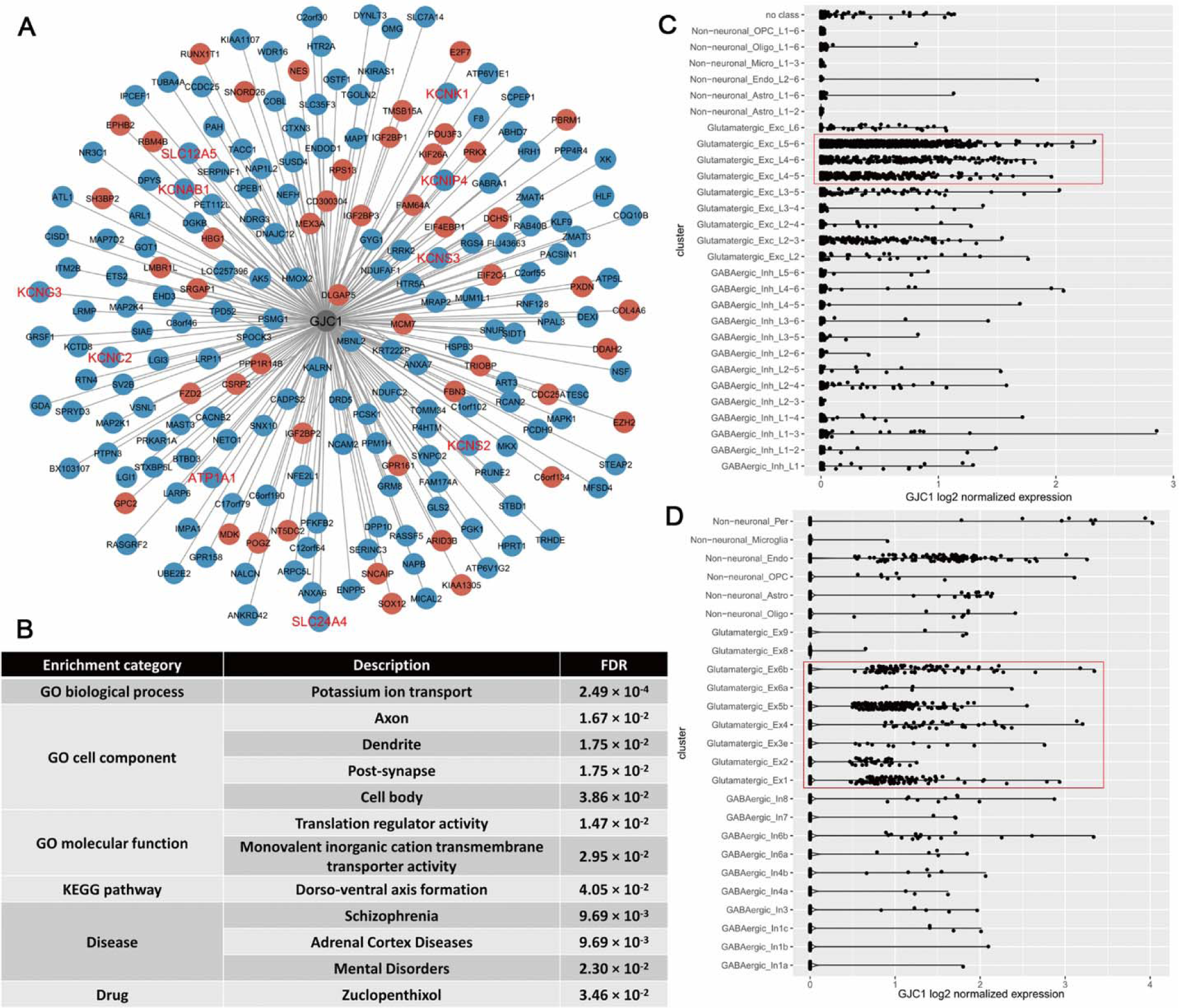
Genes co-expressed with *GJC1* were enriched in schizophrenia and schizophrenia-associated pathways and drug. A, *GJC1*-hubed co-expression network. The nodes of genes positively co-expressed with *GJC1* (*r*_*pearson*_ > 0.8) were labeled in red and the nodes of genes negatively co-expressed with *GJC1* (*r*_*pearson*_ < −0.8) were labeled in blue. The names of potassium channel genes co-expressed with *GJC1* were marked in red. B, significantly enriched terms by genes co-expressing with *GJC1*. C. Cell-type specific expression of *GJC1* in the middle temporal gyrus of human brain from Allen Brain Atlas. D. Cell-type specific expression of *GJC1* in mixed brain regions, including the frontal cortex, visual cortex and cerebellum hemisphere, from PsychENCODE.

To determine whether the expression of *GJC1* was specific to a certain cell type, we performed cell type specific analysis of *GJC1* expression. *GJC1* was predominantly expressed in glutamatergic excitatory neurons (Figure 4C and 4D, refer to Supplementary Tables S12 and S13 for pair-wise *P* values among different cell types derived from Wilcoxon rank sum test). This should be interesting as glutamatergic dysfunction has long been implicated in SCZ ^61^.

## Discussion

In this study, we used two matched SCZ-sibling family cohorts of Han Chinese origin to investigate DNMs and DNM-implicated genes in SCZ. By integrating this information with publicly available data, including brain developmental expression profiles and summary GWAS statistics, we identified DNM-implicated genes that were involved in fetal neurodevelopment. These genes may confer risks for SCZ or SCZ-associated traits, and are potential drug therapy targets for SCZ.

Connexin 45 (Cx45) encoded by *GJC1* is a component of the gap junction channel. It is one of the two connexin genes expressed predominantly in neurons ^62^. Our analyses showed that *GJC1* was expressed highly in prenatal stage as compared to that in postnatal stage and was expressed predominantly in glutamatergic excitatory neurons in the human brain (Figure 4C and 4D). For the nonsense mutation identified in our study, its occurrence might trigger the activation of surveillance pathway nonsense-mediated mRNA decay (NMD), which reduces aberrant proteins to be formed ^63^. The reduction of *GJC1* may perturb the co-expressed genes and thus influence the potassium ion transport and axon/dendrite formations, especially in early neurodevelopment (Figure 4A and 4B). Krüger *et al*. reported Cx45-deficient embryos exhibited striking abnormalities in vascular development and died between embryonic day 9.5 and 10.5 ^64^. Kumai *et al*. reported that Cx45-deficient embryos displayed an endocardial cushion defect in early cardiogenesis and died of heart failure at around embryonic day 9 ^65^. Nishii *et al*. showed that mice lacking Cx45 conditionally in cardiac myocytes displayed embryonic lethality and the requirement of Cx45 for developing cardiac myocytes ^66^. These studies indicate the critical role of Cx45 in embryonic development. Our results showed that *GJC1* displayed biased high expression across multiple brain regions in prenatal stage, suggesting it is required for neurodevelopment. The high expression of *GJC1* in prenatal/early stage in other species indicated its involvement in development is phylogenetically conserved. The dysfunction or loss-of-function can cause lethal effects, thereby such mutation would be eliminated by natural selection in evolution. Interestingly, all potassium ion channel genes were negatively co-expressed with *GJC1*. Therefore, the reduction of *GJC1* indicates an increase or overactivity of potassium channels. Miyake *et al*. reported overexpression of *Ether-a-go-go* potassium channel gene *KCNH3* in the forebrain can impair the performances of working memory, reference memory and attention ^67^. Ghelardini *et al*. reported that administrations of potassium channel openers bring out amnesic effect which can be reversed by potassium channel blockers ^68^. This evidence suggested that the cognitive or memory deficits in this case were caused by the increased expression of potassium channel genes due to the reduction of *GJC1* activity (Figure 5). The various connexins of gap junctions may be capable of differentiating between the operation qualities of the cognate synapses defined by the neurotransmitter types ^69^. Mitterauer *et al*. raised a hypothesis that if the function of glial gap junction proteins is lost, the brain is incapable of distinguishing between the same and different qualities of information processing and thus cause severe cognitive impairments in SCZ ^69^. Our findings support these hypotheses. Overall, the loss-of-function of Cx45 may be a driver of pathogenesis for some SCZ cases.

**Figure 5.**
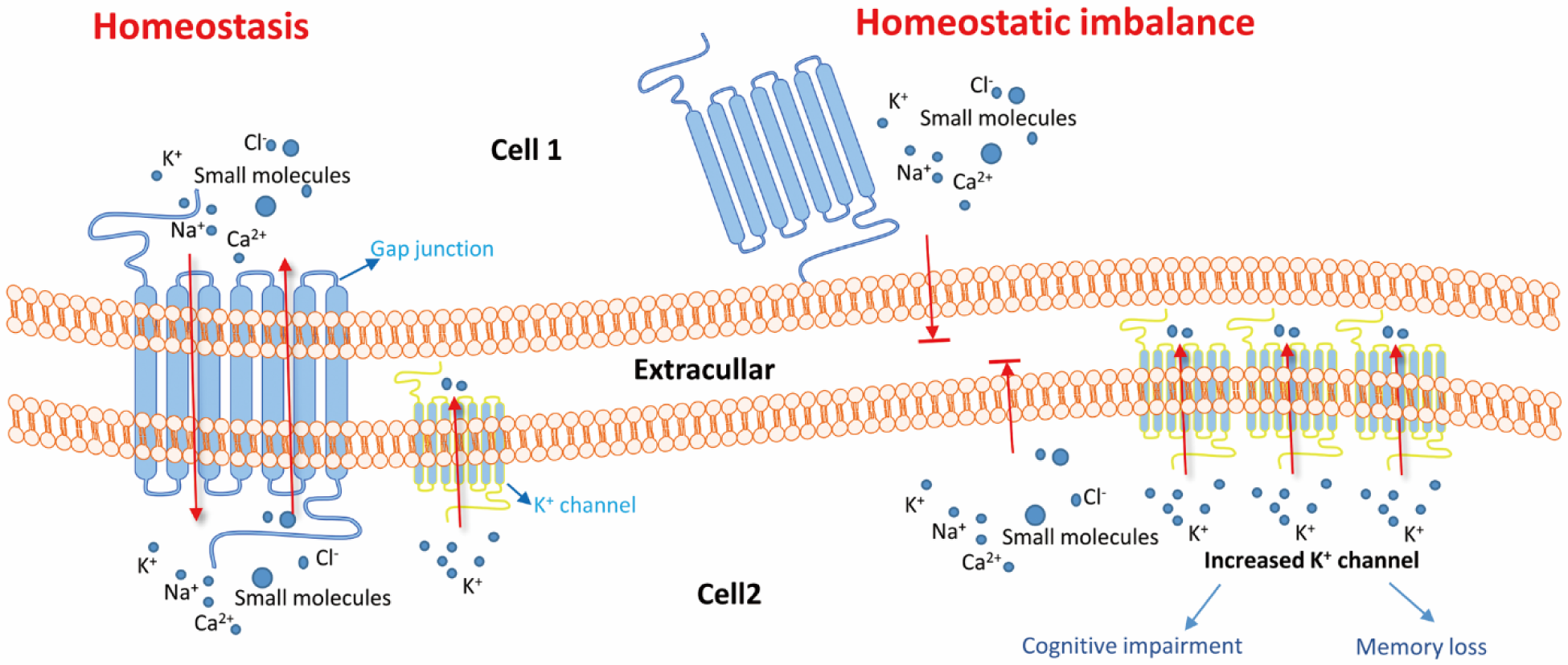
Schematic illustration of possible mechanism of dysfunction of *GJC1* leads to SCZ. Loss of gap junction formed by connexin 45 blocked the pass of ions and small molecules between brain cells, which caused the state of homeostatic imbalance in cell. Increased potassium channel subsequent to the loss of gap junction leads to cognitive impairment and memory loss in SCZ patients.

*HIST1H2AD*, the other nonsense DNM gene close to HLA locus, encodes the member D in the histone cluster 1 H2A family. Studies have demonstrated that several SCZ candidate genes are especially susceptible to changes in transcriptional activity as a result of histone modification ^70, 71^. Therefore, a deficit in the histone itself can implicate multiple SCZ candidate genes. Moreover, epigenetic regulation effects of histone deacetylase inhibitors were potentially suggested to treat SCZ ^72^, which implies the fundamental role of histone in the pathology of SCZ. Gene level DNM burden test identified a list of genes carrying more DNMs in SCZ patients than their unaffected siblings. In the literature, there were experimental or clinical evidences demonstrating their potential association with SCZ. Luan et al. reported SNPs in *MSI2* are strongly associated with SCZ in the Chinese population ^73^. In the GWAS of PGC, the SNPs in *MSI2* were also associated with SCZ (Supplementary Table 7). Another DNM-burdened genes, *DPYSL2*, is a member of the collapsin response mediator protein (CRMP) family. CRMP forms homo- and hetero-tetramers and facilitates neuron guidance, growth and polarity. It also plays a role in synaptic signaling through interactions with calcium channels. Lee *et al*. revealed that *DPYSL2* was downregulated in the PFC and hippocampus of prenatally stressed (PNS) adult rats that underperformed in behavioral tests ^74^. Bruce *et al*. conducted potassium channel-targeted SNP association analyses with SCZ and SCZ-associated phenotypes. rs8234 in *KCNQ1*, a DNM-burdened gene in our study, was associated to processing speed ^75^. Geschwind *et al*. found that patients with voltage-gated potassium channel complex antibody (VGKCC-Abs) had particular impairment in memory and executive functions when they evaluated cognitive function and imaging data in patients with VGKCC-Abs associated encephalopathy ^76^. Interestingly, the impaired brain functions coincide with the two most implicated brain regions in SCZ, dorsolateral prefrontal cortex and the hippocampus ^77^. This evidence demonstrated the efficacy of our DNM burden test by incorporating enhancer and promoter regions into consideration. It also indicated DNMs contributing to the genesis of disease are not limited in coding regions, underling the value of WGS.

Except for *HIST1H2AD*, common variants in these identified DNM genes were not strongly associated to psychiatric diseases. However, a few of them, *GJC1, HIST1H2AD* and *SDK1*, were strongly associated to SCZ-associated traits, including intelligence, educational attainment, smoking and drinking. It suggested the significance of these DNM genes in neurodevelopment and that the lethal mutations occurred in these genes were eliminated by marked reduction of fecundity in psychiatric patients.

The results should be interpreted with caution because of limited samples in the pilot study. A few parents with other mental illness from the Taiwan cohort may complicate the interpretation of the DNMs if the mental illness shares certain genetic liability with SCZ. Nevertheless, the association of the nonsense DNM gene *GJC1* with SCZ is deemed reliable: First, the association of *GJC1* with SCZ is supported by the analyses of rare variants of an independent exome sequencing dataset where SCZ patients have more rare variants in the gene than controls^27^. Second, with its intolerance of loss-of-function index of pLI = 0.94, the loss-of-function mutation is very likely to have detrimental consequences in the regulation of ion channels which have been implicated in the pathogenesis of SCZ^78, 79^. To validate our findings, future studies with more and independent Chinese family samples are necessary.

In summary, we identified a list of DNM-implicated genes which are involved in prenatal neurodevelopment. Common variants in these DNM-implicated genes had been previously reported to be associated with SCZ and related traits in previous GWASs, which is consistent with our analyses. DNMs implicating the enhancers may contribute to pathogenesis of SCZ by dysregulating the expression of genes. *GJC1* is one of these DNM-implicated genes that is primarily expressed in glutamatergic neurons and may be involved in the modulation of ion channel functions, which has also been implicated in SCZ in previous studies ^78, 79^. Overall, our study provided new evidence that DNMs have a significant role in SCZ. Further study of these DNM-implicated genes with functional analyses could lead to better understanding of the pathology of SCZ.

## Data Availability

The whole genome sequencing data used in the manuscript can be accessed from NCBI.

https://www.ncbi.nlm.nih.gov/bioproject/PRJNA551447

## Acknowledgements

This work was supported in part by grants from National Institutes of Health (R01MH101054 to X.C. and R01LM012806 to Z.Z.), the National Natural Science Foundation of China (grant 81421061), the National Key Research and Development Program (2016YFC0906400), Shanghai Key Laboratory of Psychotic Disorders (13dz2260500), Cancer Prevention and Research Institute of Texas (RR180012 to X.J.), and UT Stars award to X.J. The DNA samples of the subjects from Taiwan cohort were obtained through NIMH Genetics Repository. The DNA samples of the subjects from Shanghai cohort were from the Bio-x SCZ Biobank in Shanghai, China. Computational resources from the school of biomedical informatics at The University of Texas Health Science Center at Houston were used in data analysis. The data from the TSLS were collected with funding from grant R01MH59624 from K. S. K. and M. T. The whole genome sequencing of samples from Taiwan cohort were supported by grant from National Institutes of Mental Health (1RO1-MH085560) to M.T. We acknowledge Dr. Hai-Gwo Hwu and Dr. Wei J. Chen for their recruitment of families, collection of clinical data and preprocess of blood samples in Taiwan cohort. We acknowledge the help from Dr. Lukas Simon on single cell analysis and the collection of GWAS summary statistics from Dr. Yulin Dai. We acknowledge the altruism of the participants and their families and support staff at each of the participating sites for their contributions to this study.

## Author contributions

S.J. designed the study, performed the analyses, interpreted the results and wrote the manuscript. D.Z. collected the demographic data and performed whole genome sequencing, data processing, quality control and cleaning from Shanghai cohort. Y.W., P.J., C.W., X.L., G.H., D.C., X.J., T.M. and J.S.W. contributed to data processing, quality control and cleaning. K.S.K. and M.T. contributed to data collection and whole genome sequencing. J.S.W., Y.L. and J.C. conducted Sanger sequencing experiment. T.M. revised the manuscript. L.H., J.C., Z.Z., P.J. and X.C. conceived the project, designed the study, collected the data, interpreted the results and wrote the manuscript.

